# Longitudinal pathways between maternal depression, parenting behaviors, and early childhood development: a mediation analysis

**DOI:** 10.1101/2024.01.24.24301747

**Authors:** Allison Frost, Elissa Scherer, Esther O. Chung, John A. Gallis, Kate Sanborn, Yunji Zhou, Ashley Hagaman, Katherine LeMasters, Siham Sikander, Elizabeth Turner, Joanna Maselko

## Abstract

Maternal depression is a global public health concern with far-reaching impacts on child development, yet our understanding of mechanisms remains incomplete. This study examined whether parenting mediates the association between maternal depression and child outcomes. Participants included 841 rural Pakistani mother-child dyads (50% female). Maternal depression was measured at 12 months postpartum, parenting behaviors (warmth, stimulation, and harsh parenting) were measured at 24 months, and child outcomes (mental health, socioemotional development, and cognitive skills) were measured at 36 months. Maternal depression predicted increased harsh parenting, child mental health difficulties, and child socioemotional concerns; however, there was little evidence for parenting as a mediator between maternal depression and child outcomes. Sex-stratified results are discussed, and findings are situated in context.

---

Extensive evidence indicates that maternal depression has intergenerational impacts on the physical, cognitive, and socioemotional development of offspring (Avan et al., 2010; Goodman et al., 2011; Patel et al., 2004; Stein et al., 2014; Surkan et al., 2012). Infants of mothers experiencing depression are known to have significantly lower height and weight, social and emotional development scores, motor development abilities, greater attachment insecurity, and delayed language development, among other adverse outcomes (Gelaye et al., 2016). Globally, the prevalence of maternal depression is high, with estimates in low and middle income countries (LMICs) reaching 50% (Parsons et al., 2012). Maternal depression is a major public health priority for improving child development.

Interventions targeting maternal depression alone show limited impacts on child development (Pitchik et al., 2020). Therefore, it is important to understand the mechanisms linking maternal depression and child outcomes that can serve as promising targets for interventions (Goodman & Garber, 2017). An integrative model of maternal depression and child development proposed by Goodman and Glotlib (1999) posits that there are many complex pathways linking maternal depression to child adverse outcomes, with overlapping mediators (e.g., genetics, epigenetics, neuroendocrine dysregulation, parenting) and moderators (e.g., child temperament, family stress) (Goodman & Gotlib, 1999; Goodman et al., 2020). Among these, parenting is an especially promising area of focus for interventions, as there is a strong theoretical body of work examining how parenting is impacted by maternal depression and parenting behaviors are modifiable (Goodman et al., 2020).

Parenting is a multifaceted construct comprised of several domains, including parental warmth and responsiveness (i.e., the extent to which the parent is attentive to the child’s needs and responds in an affectionate manner), discipline and control (i.e., the parent’s strategies for managing their child’s behavior), and scaffolding and stimulation (i.e., behaviors to structure the child’s environment and encourage learning and development) (Culpin et al., 2020). Parent warmth and responsiveness (Eshel et al., 2006; Scherer et al., 2019) and parent scaffolding and stimulation behaviors (Obradović et al., 2016) are known to support children’s socioemotional and cognitive development. Furthermore, harsh parenting practices (i.e., interactions with the child that involve high amounts of hostility, emotional reactivity, or physical discipline) are associated with mental health difficulties and poor socioemotional development in children (McCullough & Shaffer, 2014; Speyer et al., 2022; Wolford et al., 2019).

The action-control framework proposed by Dix and colleagues (2009) postulates that depression influences parenting through several co-occurring pathways, such as limited maternal attention toward their children, negative appraisals of child behavior, increased displays of negative emotions, and decreased displays of positive emotions during mother-child interactions (Dix & Meunier, 2009). Together, these processes may interfere with mothers’ ability to exhibit warmth and responsiveness toward their children (Goodman et al., 2017; Madeghe et al., 2016; Unternaehrer et al., 2019). Mothers with depression may also be more likely to exhibit negative affect or withdrawn behavior towards the child, or to choose harsh punishment strategies when the child misbehaves (McCabe, 2014; Vreeland et al., 2019; Wolford et al., 2019). It is important to note that several other factors are also known to influence maternal parenting behaviors including maternal social support (Thomson et al., 2014), wealth (Vreeland et al., 2019), previous life experiences such as adverse childhood experiences (ACEs) (Lange et al., 2019), education (Thomson et al., 2014; Unternaehrer et al., 2019; Vreeland et al., 2019; Woodward et al., 2018), and age (Thomson et al., 2014)

A recent systematic review by Goodman and colleagues concluded that there was strong empirical evidence for parenting mediating the relationship between maternal depression and child outcomes (Goodman et al., 2020). However, there are several studies showing that parenting does not always mediate the association between maternal depression and child development (Goodman et al., 2020; Gueron-Sela et al., 2018; McManus & Poehlmann, 2012). Existing papers persistently suggest that the associations among maternal depression, parenting, and child development are highly dependent on context. For instance, studies in Asian cultures have drawn attention to supportive postpartum rituals that can be protective for both mother and child outcomes (Gulamani et al., 2013; LeMasters et al., 2020). However, the vast majority of research in this area has occurred in relatively homogenous contexts, namely North American and European countries ( Goodman et al., 2020). More research is needed across diverse contexts, including Asian countries and low-resource settings (such as LMICs), to build the evidence base around maternal depression and child outcomes.

Few studies in low-resource, Asian settings have tested whether parenting mediates the association between maternal depression and child development outcomes. For instance, one study in rural Bangladesh by Black and colleagues (2007) found that maternal and paternal responsiveness and stimulation partially mediate the relationship between maternal depression and cognitive development in infants (Black et al., 2007). Another study by Tran and colleagues in Vietnam (2014) found that antenatal depression was not directly associated with child socioemotional development, but the relationship was mediated through parenting practices during infancy, including lower affection and warmth (Tran et al., 2014). Sparse research on this topic in these settings is additionally limited by lack of longitudinal data. An important next step in expanding this literature is establishing the context specific, longitudinal associations among maternal depression, parenting, and child outcomes.

In Pakistan, there are several unique contextual factors that may influence the associations among maternal depression, parenting, and child development. Households in this region are likely to include extended families (rather than nuclear families), providing both additional childcare support and potential increased conflict between family members (Aubel, 2012; Chung et al., 2022; Chung et al., 2020). In addition, family relationships are emphasized in this context. This may provide a sense of belonging and familial responsibility among children; however, such interrelatedness may also increase children’s sense of shame, as any misbehavior is thought to reflect poorly on both the child and their family (Zaman, 2014). Pakistan is also a highly patriarchal culture, where children are likely to be treated differently based on their sex.

For instance, parents may be more likely to encourage autonomy in boys compared to girls; however, parents may show more warmth to girls compared to boys (Stewart et al., 2006). Finally, additional stressors such as poverty, exposure to intimate partner violence (IPV), and lack of access to resources are salient to both mother and child experiences (Hagaman et al., 2020; Haight et al., 2022; Maselko et al., 2018).

This study aimed to investigate if maternal warmth, stimulation, and harsh parenting behaviors mediated relationships between maternal depression and the mental health, socioemotional, and cognitive development of children in rural Pakistan over time. We also explore differences in these associations by child sex. Based on previous literature, we hypothesize that maternal parenting behaviors will mediate the association between maternal depression and child outcomes. Specifically, mothers with depression will show lower warmth, lower stimulation behaviors, and higher harsh parenting behaviors, which will then predict increased child mental health difficulties, increased socioemotional problems, and decreased cognitive skills. This study expands on limited literature in this context, providing a longitudinal examination of parenting and child development.

## Method

### Study Sample

Data for these analyses are from the Bachpan cohort study, a population-representative longitudinal birth cohort established in the context of a perinatal depression intervention in rural Pakistan. The full study details have been published elsewhere (Turner et al., 2016). Briefly, all women in their 3^rd^ trimester of pregnancy residing within the study area were screened for depression using the Patient Health Questionnaire-9 (Kroenke et al., 2001). Those who scored 10 or greater on the PHQ-9 (met screening criteria for likely depression) were invited to join the study. Additionally, women who scored less than 10 (did not meet screening criteria for depression) were invited to create balanced sample sizes of depressed and non-depressed women. Additional inclusion criteria were that the women were married, understood one of the languages the study was carried out in (Urdu Punjabi, or Potohari), were planning on staying within the study area, and were not in need of any immediate medical attention. In total, 1,154 women were enrolled in the study at baseline (n = 283 intervention, n = 287 control, n = 584 non-depressed). For this analysis, we used data from interviews conducted at pregnancy (baseline), and 12, 24, and 36 months post-partum. The analytic sample includes 841 dyads. The present study examined maternal depression at 12 months postpartum, parenting at 24 months, and child outcomes at 36 months. Intervention or control arm random assignment was based on the village cluster where each woman resided. Trial arm was included as a covariate in all analyses.

## Measures

### Maternal Depression

Maternal depression was measured at child age 12 months using the Structured Clinical Interview for the DSM-IV (SCID) Major Depression Module (First et al., 2002). The SCID is a semi-structured psychiatric interview. Assessors determined the presence of a current Major Depressive Episode based on participant symptoms, resulting in a binary indicator of depression status. The SCID has been adapted for use with postpartum women and was translated into Urdu and adapted for use in Pakistan by the study team (Gorman et al., 2004; Rahman et al., 2003).

### Parenting

The Observation of Mother-Child Interaction (OMCI) was used to assess parenting behaviors at the 24-month timepoint. The OMCI is a tool used to directly observe and code responsive caregiving interactions during a structured activity between the caregiver and child. The tool was created in Pakistan to allow for direct observation of caregiving behaviors without the use of video (Rasheed & Yousafzai, 2015; Scherer et al., 2019). The OMCI has shown cross-sectional associations with maternal depression, child cognitive development, and other observational measures of parenting behavior (Rasheed & Yousafzai, 2015; Scherer et al., 2019). Within the context of this study, the interaction consisted of a 5-minute observation of the mother and child interacting with a picture book. Before beginning the activity, the mother was told that the observer wanted to watch her talk and play with her child with the picture book.

Throughout the interaction, the observer coded 11 maternal behaviors. Observed behaviors centered around maternal warmth (e.g., positive touch, positive verbal statements), stimulation (e.g., asking the child questions, pointing to items in the book), and harsh parenting (e.g., negative affect, negative verbal statements). Behaviors are scored based on the frequency of their occurrence (i.e. 0 = never occurred, 1= occurred 1-2 times, 2= occurred 3-4 times, or 3 = occurred 5+ times) and items are averaged to create a summary score for each domain.

### Child outcomes

All child outcomes were assessed when children were 36 months old.

### Strengths and Difficulties Questionnaire (SDQ)

The Strengths and Difficulties Questionnaire (SDQ) was used to assess mental health difficulties (Goodman, 1997; Samad et al., 2005). The SDQ is a parent-reported questionnaire comprised of 25 questions on child mental health concerns, including anxiety and depressive symptoms, conduct problems, hyperactivity, and problems with peers. Parents rate each item on a scale from 0 (“never true”) to 2 (“certainly true”) and items are summed to create a Total Difficulties score (Range: 0-40; α = 0.78), with higher scores being indicative of greater difficulties. The SDQ also includes subscales for each type of difficulty (emotional problems, conduct problems, hyperactivity problems, and peer problems) and a prosocial scale, which is not included in the total mental health difficulties summary score.

### Ages and Stages Questionnaire – Socioemotional (ASQ-SE)

The Ages and Stages Questionnaire (ASQ-SE) was used to assess socioemotional difficulties (Juneja et al., 2012; Squires et al., 2001). The ASQ-SE assesses the frequency of developmentally appropriate behaviors. The instrument is comprised of 30 questions that are answered by a caregiver. For each question (e.g., does your child laugh or smile when you play with him/her?), a point value is assigned based on the responses (i.e., “most of the time” = 0 points [or 10 points if reverse-coded], “sometimes” = 5 points, “rarely or never” = 10 points [or 0 points]). Items are summed to create an overall socioemotional difficulties score (α = 0.65). The range is 0-300 where the higher the score, the more cause for concern about the child’s behaviors.

### Bayley scales

Two of the five Bayley Scales of Infant and Toddler Development (receptive language and fine motor) were used to assess cognitive outcomes (Bayley & Reuner, 2007). The receptive language scale assesses the child’s ability to understand and respond to the words of the assessor. The fine motor scale is used to assess the child’s ability to manipulate objects and their hand function. The scales were administrated by a trained assessor. Following this, scaled scores were calculated using the child’s chronological age. Scores range from 1 to 19, with higher scores indicating increased language and motor skills.

### Other Variables of Interest

Additional variables were identified as potential confounders based on the existing literature and a directed acyclic graph. These included maternal and paternal education (i.e., grades passed) at baseline (Davis-Kean et al., 2019), family structure at baseline (nuclear v. extended) (Allendorf, 2013), whether the mother experienced interpersonal violence (IPV) in the last 12 months (measured during the 12 month visit) (Neamah et al., 2018), the total number of adverse childhood experiences (ACEs) the mother had experienced (measured at the 36-month visit) (Sun et al., 2017), and household assets at baseline (Hamadani et al., 2014). The household asset score was a composite score comprised of items such as whether the family owns land, the type of material used to build their housing, and whether the family owns certain household items (e.g., a television) ( Maselko et al., 2018). A polychoric principal components approach was used to derive an asset index from these items, resulting in a single continuous asset score (Mean = 0.00, Range = −5.04, 2.84) that can be used as an indicator of socioeconomic status ( Maselko et al., 2018). Finally, trial arm (intervention, control, non-depressed at baseline) was included as a covariate. The intervention was not significantly associated with any outcomes in this study (Maselko et al., 2020).

## Statistical Methods

Structural equation modeling (SEM) was used to test direct and indirect pathways from maternal depression to child outcomes (Madeghe et al.). The primary predictor was maternal depression, a binary manifest variable. Latent factors were created for each domain of parenting (warmth, stimulation, and harsh parenting) and child mental health (SDQ) and socioemotional (ASQ-SE) outcomes. Child cognitive outcomes (Bayley’s fine motor and receptive language scores) were modeled as scaled scores. Model fit was evaluated using the Comparative Fit Index (CFI), Tucker-Lewis Index (TLI) and Root Mean Square of Approximation (RMSEA). CFI and TLI values over 0.9 and RMSEA values under 0.05 are indicative of good model fit (Hu & Bentler, 1999) and modification indices were used to improve model fit as needed. In addition, we used full information maximum likelihood to account for missing data (Enders & Bandalos, 2001).

We tested the direct effect of maternal depression at 12 months on all child outcomes (SDQ, ASQ-SE, Bayley fine motor, Bayley receptive language) at 36 months, as well as the indirect effects of maternal depression on child outcomes through parental warmth, stimulation, and harsh parenting at 24 months. Bootstrapped confidence intervals were computed for indirect effects using 1,000 bootstrap resamples in two of the four outcomes (SDQ and Bayley fine motor). For the ASQ and Bayley receptive language, mediation results were not bootstrapped due to issues with convergence. In additional analyses, we test for sex differences in observed effects by stratifying models by child sex.

For each outcome, we also ran a set of sensitivity analyses. First, we control for assessors at 12, 24, and 36 months. Then, we further investigate missing data in our exogenous variables. Specifically, we impute missing data for IPV exposure at 12 months and ACEs exposure at 36 months. We test each model with the “best case scenario”, meaning all women with missing data on IPV were assigned a zero (i.e., no IPV experienced) and all women with missing ACEs data were assigned zero (i.e., no adverse childhood experiences). We also test the “worst case scenario” in which all women with missing IPV data were assigned a one (i.e., experienced IPV) and all women with missing ACEs data were assigned the median ACEs score (Median = 1.00)

## Results

Participants in this study included 841 mother-child dyads (Table 1). On average, mothers were 26.68 years old at baseline and completed 7.76 years of education. Mothers had 1.45 living children on average at baseline. 87.28% of mothers lived in a household with extended family and 18.21% of mothers met criteria for depression at 12 months postpartum. Children were 50.3% male and 49.7% female.

**Table 1.**
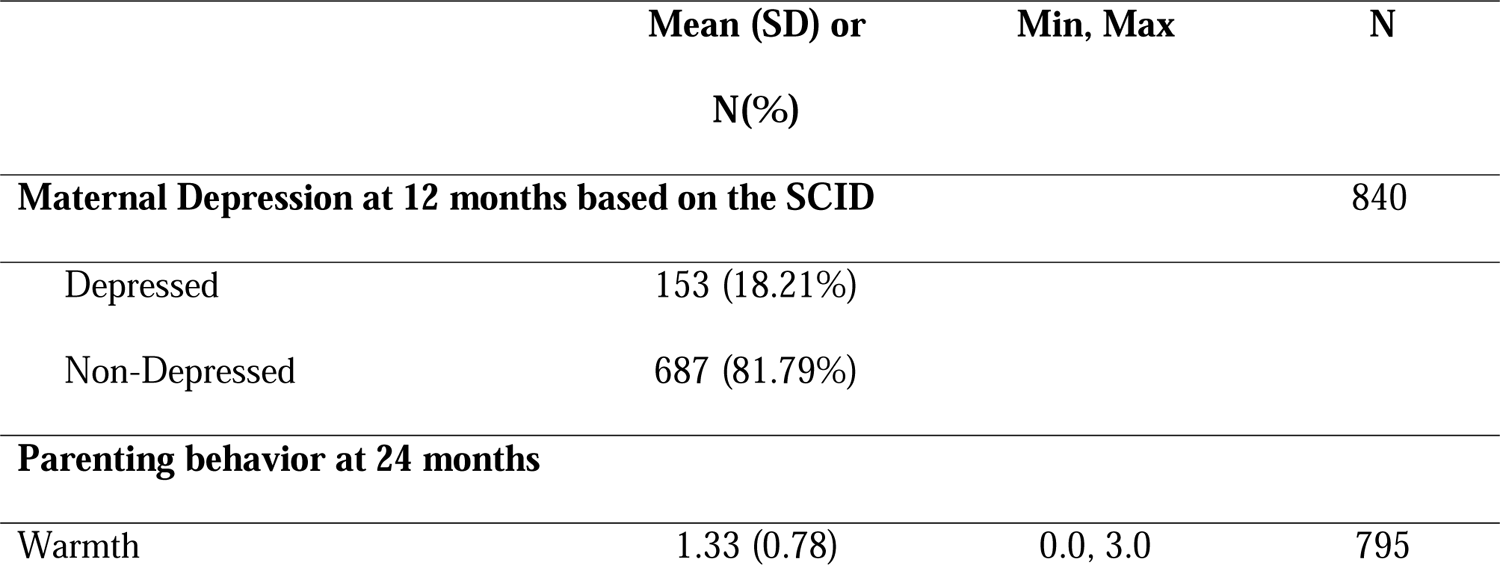
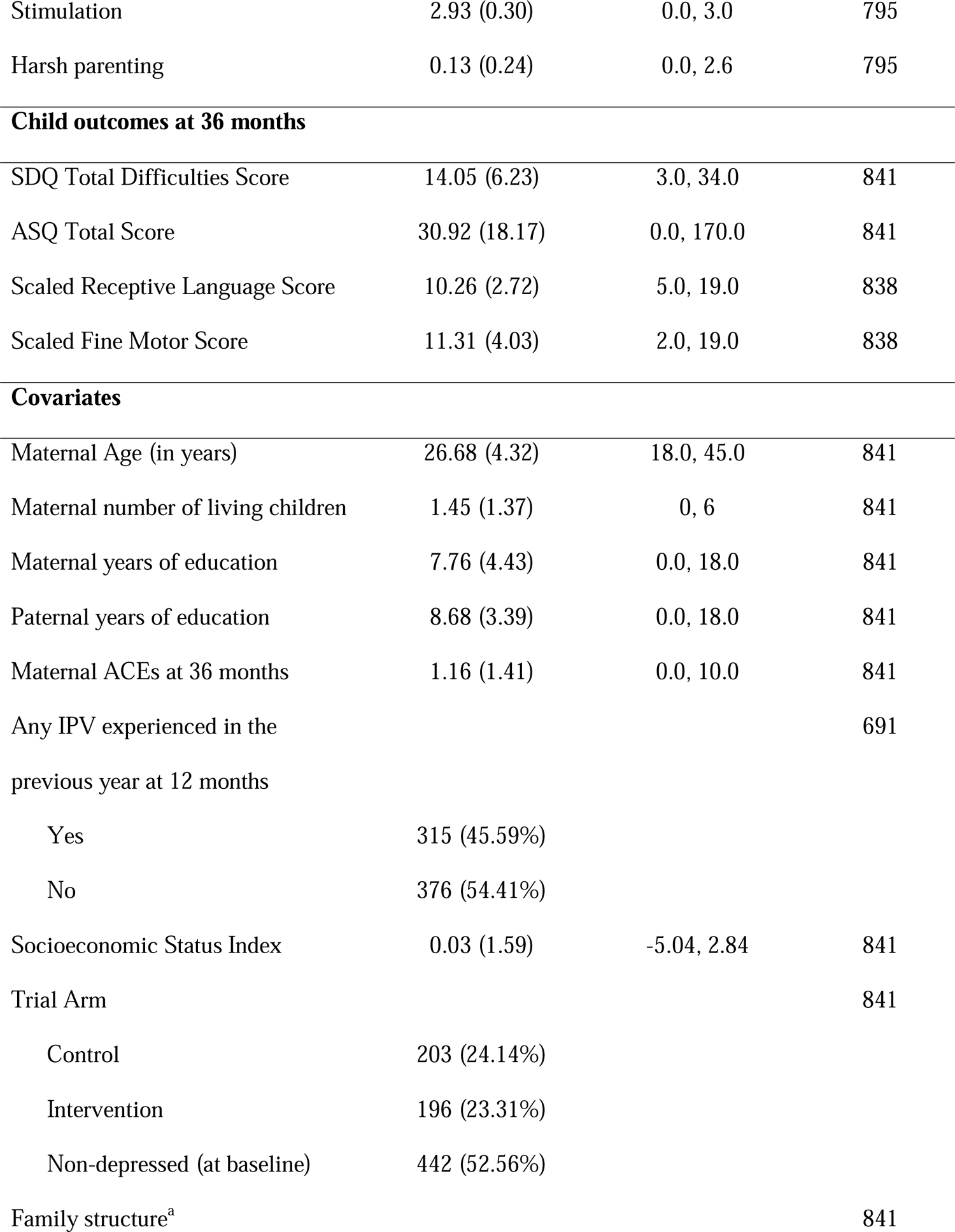
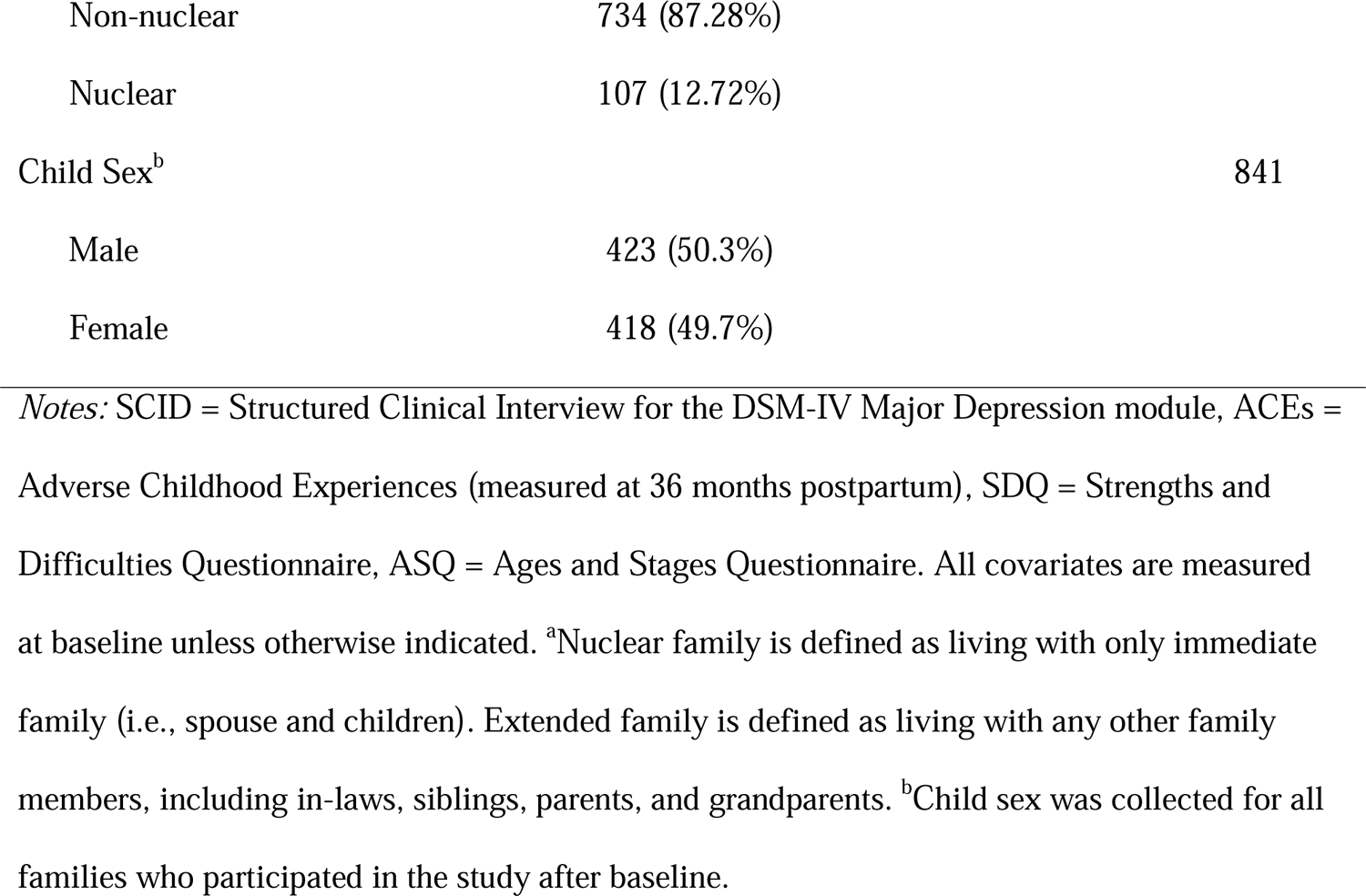
Descriptive statistics (n=841)

Results of the structural equation modeling are described by outcome below. Latent factors for maternal warmth, stimulation, and harsh parenting were created. The measurement model for the parenting factors was kept constant across all outcomes. For each outcome, we present the direct effects of maternal depression on child outcomes and parenting behaviors, the direct effects of parenting behaviors on child outcomes, and the indirect effects of maternal depression on child outcomes through parenting behaviors. We then present results stratified by child sex. All presented effects are standardized.

### Strengths and Difficulties Questionnaire (SDQ)

Model results are presented in Figure 1. We created a latent factor representing child mental health difficulties using items from the SDQ. To improve model fit, redundant indicators based on modification indices were removed from the SDQ factor and a subset of items were retained (see Appendix A for included items). In addition, based on modification indices, the OMCI item measuring maternal positive affect was loaded on to the Harsh Parenting factor. Model fit was as follows: CFI: 0.88, TLI: 0.87, RMSEA: 0.04. There was a significant direct effect of maternal depression on child SDQ (β = 0.39, 95% CI: 0.18, 0.59), suggesting that children whose mothers were depressed at 12 months postpartum showed increased mental health difficulties at 36 months, after adjusting for potential confounders. There was also a significant association between maternal depression at 12 months and harsh parenting at 24 months (β = 0.43, 95% CI: 0.15, 0.71), suggesting that mothers with depression showed increased harsh parenting behaviors. Maternal depression was not significantly associated with maternal warmth or stimulation. In addition, maternal warmth, stimulation, and harsh parenting were not significantly associated with child SDQ. Finally, there were no statistically significant indirect effects from maternal depression to child SDQ through any parenting factor.

**Figure 1.**
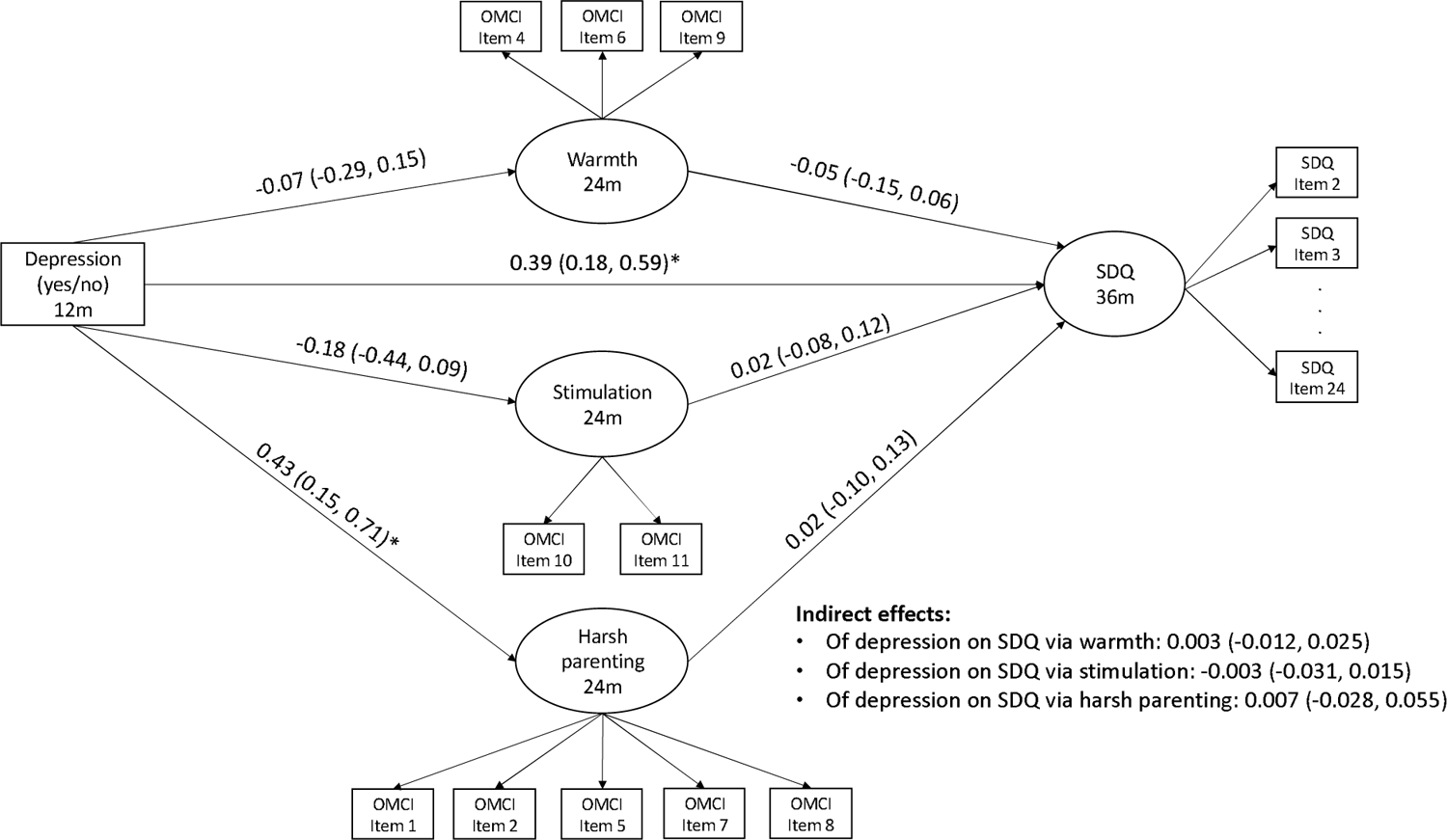
Structural equation model with maternal depression at 12 months predicting child mental health difficulties at 36 months through parenting at 24 months. Notes: Standardized effect sizes with 95% confidence intervals (CIs) are presented. Indirect effect estimates include bootstrapped CIs. Model controls for maternal adverse childhood experiences, past year intimate partner violence exposure, maternal education, paternal education, socioeconomic status, family structure, and trial arm. Model includes correlations among all parenting items. SDQ = Strengths and Difficulties Questionnaire, OMCI = Observation of Mother Child Interaction *p < .05

Results stratified by child sex are presented in Supplemental Table 1. The association between maternal depression and harsh parenting was significant for girls (β = 0.50, 95% CI: 0.08, 0.92), but not boys (β = 0.30, 95% CI: −0.75, 1.34). All other effects were similar between sexes. As a secondary analysis, we examined each subscale of the SDQ (emotional problems, conduct problems, hyperactivity problems, peer problems, and prosocial behavior). Results are presented in Supplemental Table 2. We found that maternal depression was associated with increased emotional problems (β = 0.34, 95% CI: 0.11, 0.56), conduct problems (β = 0.39, 95% CI: 0.18, 0.60), and peer problems (β = 0.22, 95% CI: 0.04, 0.40). There were no significant associations between maternal depression and hyperactivity or prosocial behavior. Maternal warmth was negatively associated with emotional problems, such that children whose mothers showed more warmth showed lower emotional problems (β = −0.11, 95% CI: −0.21, −0.01). There were no other associations between parenting behaviors and SDQ subscales, nor were there indirect effects from maternal depression to SDQ subscales through parenting. Adjusting for assessors (see Supplemental Table 3) and sensitivity analyses with IPV and ACEs (see Supplemental Table 4) yielded similar results.

### Ages and Stages Questionnaire-Socioemotional (ASQ-SE)

Figure 2 shows the ASQ-SE model results. The model fit was as follows: CFI: 0.82, TLI: 0.80, RMSEA: 0.04. As with the SDQ model, modification indices were used to remove redundant ASQ-SE indicators as well as items with low variability (see Appendix A for a list of included items). There was a significant direct effect from maternal depression at 12 months to child ASQ-SE at 36 months (β = 0.27, 95% CI: 0.08, 0.47). Mothers with depression at 12 months reported increased ASQ-SE scores for their children at 36 months, indicating increased concerns around child socioemotional development. In addition, as with the SDQ model, depression was associated with increased harsh parenting. There were no other significant associations between depression and parenting, nor were there significant associations between parenting and ASQ-SE scores. Finally, there were no significant indirect effects of maternal depression on child ASQ-SE scores through maternal warmth, stimulation, or harsh parenting.

**Figure 2.**
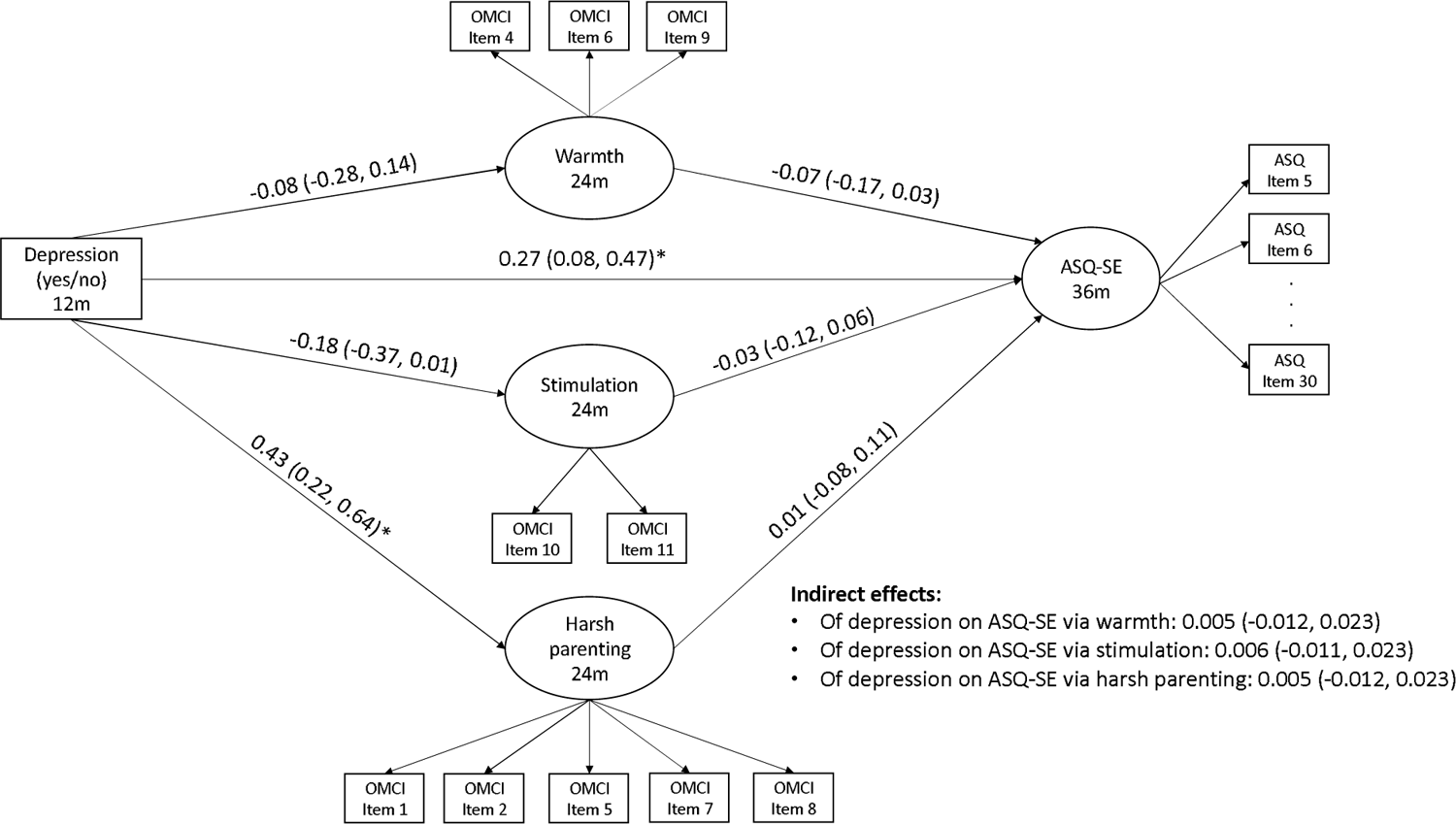
Structural equation model with maternal depression at 12 months predicting child socioemotional development at 36 months through parenting at 24 months. Notes: Standardized effect sizes with 95% confidence intervals (CIs) are presented. Model controls for maternal adverse childhood experiences, past year intimate partner violence exposure, maternal education, paternal education, socioeconomic status, family structure, and trial arm. Model includes correlations among all parenting items. ASQ-SE = Ages and Stages Questionnaire-Socioemotional Development (higher scores indicate increased concerns), OMCI = Observation of Mother Child Interaction *p < .05

Model results by child sex are presented in Supplemental Table 5. The association between maternal depression and child ASQ-SE was significant for boys (β = 0.40, 95% CI: 0.13, 0.68), but not girls (β = 0.16, 95% CI: −0.12, 0.44). Similar to the SDQ model, we found that the association between maternal depression and harsh parenting was stronger for girls compared to boys. There were no other model differences by child sex.

As a sensitivity analysis, we also tested this model with ASQ-SE scores modelled as a manifest variable (i.e., the total summed ASQ score). This model had improved fit (CFI: 0.91, TLI: 0.89, RMSEA: 0.04). Model results (presented in Supplemental Table 6) were similar, although in the manifest model, harsh parenting was significantly associated with ASQ-SE scores (β = 0.09, 95% CI: 0.003, 0.17). In addition, we found similar results when adjusting for assessor (see Supplemental Table 7) and analyzing missing data on IPV and ACEs (See Supplemental Table 8).

### Bayley Scales of Infant Development – Receptive Language score

Model results are presented in Figure 3. Model fit was: CFI: 0.89, TLI: 0.86, RMSEA: 0.05. The direct effect of maternal depression on child receptive language was null. Maternal depression predicted increased harsh parenting, but not warmth or stimulation. There were no significant effects of parenting on receptive language, nor were there indirect effects of maternal depression on language through parenting.

**Figure 3.**
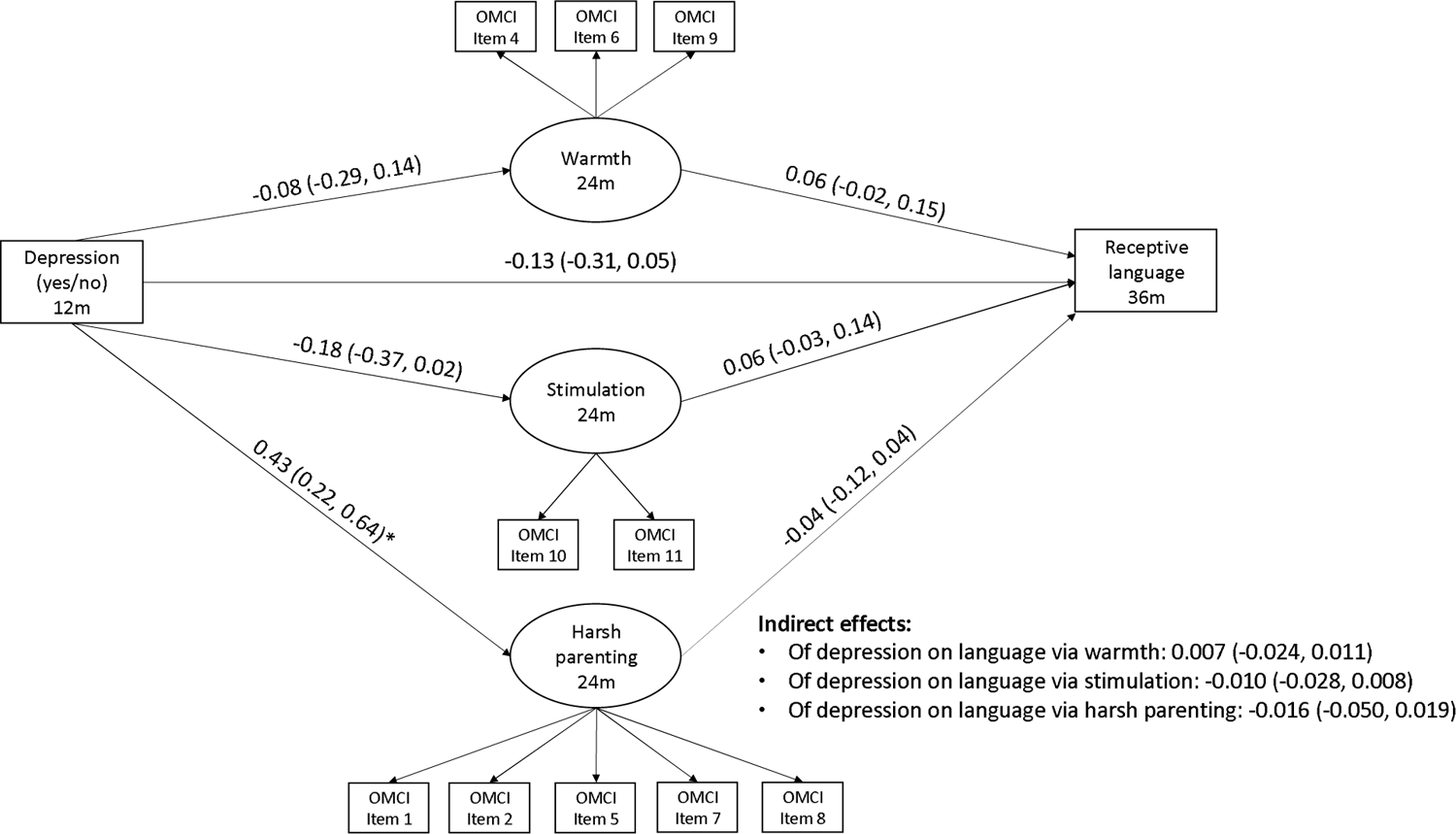
Structural equation model with maternal depression at 12 months predicting child receptive language at 36 months through parenting at 24 months. Notes: Standardized effect sizes with 95% confidence intervals (CIs) are presented. Model controls for maternal adverse childhood experiences, past year intimate partner violence exposure, maternal education, paternal education, socioeconomic status, family structure, and trial arm. Model includes correlations among all parenting items. OMCI = Observation of Mother Child Interaction *p < .05

Results stratified by child sex are presented in Supplemental Table 8. As with earlier models, the association between maternal depression and harsh parenting was stronger for girls compared to boys. In addition, the association between maternal depression and language was stronger for boys (β = −0.25, 95% CI: −0.51, 0.01) compared to girls (β = −0.01, 95% CI: −0.25, 0.24), as was the association between maternal stimulation and language (boys: β = 0.13, 95% CI: 0.00, 0.26; girls: β = 0.01, 95% CI: −0.10, 0.12). However, these associations were not statistically significant for either sex. All other model effects were similar between sexes. Adjusting for assessors at 12, 24, and 36 months (see Supplemental Table 9) and investigating missing data on IPV and ACEs (See Supplemental Table 10) yielded similar results.

### Bayley Scales of Infant Development-Fine Motor score

Figure 4 shows model results for the Bayley fine motor score. Model fit was as follows: CFI: 0.89, TLI: 0.87, RMSEA: 0.05. There was no significant direct effect of maternal depression at 12 months on child fine motor skills at 36 months. As with previous models, maternal depression at 12 months was associated with increased harsh parenting at 24 months. In addition, harsh parenting at 24 months was associated with decreased fine motor skills at 36 months (β = −0.12, 95% CI: −0.21, −0.02). There was a small, significant indirect of maternal depression on fine motor skills through harsh parenting, in which maternal depression at 12 months predicted increased harsh parenting at 24 months, which then predicted decreased fine motor skills at 36 months (β = −0.052, 95% CI: −0.109, −0.007). There were no other significant associations.

**Figure 4.**
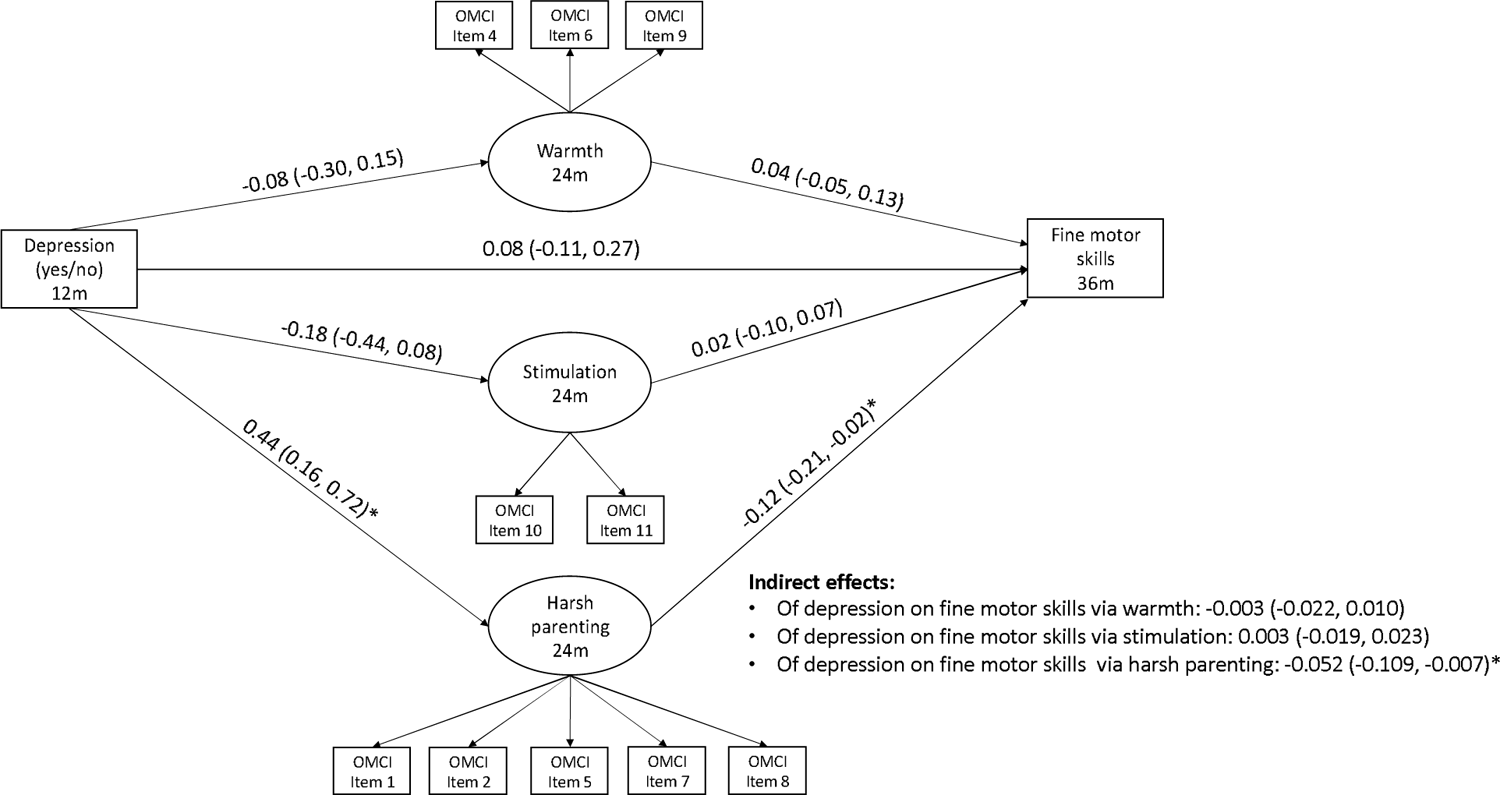
Structural equation model with maternal depression at 12 months predicting child fine motor skills at 36 months through parenting at 24 months. Notes: Standardized effect sizes with 95% confidence intervals (CIs) are presented. Indirect effect estimates include bootstrapped CIs. Model controls for maternal adverse childhood experiences, past year intimate partner violence exposure, maternal education, paternal education, socioeconomic status, family structure, and trial arm. Model includes correlations among all parenting items. OMCI = Observation of Mother Child Interaction *p < .05

Results stratified by child sex are presented in Supplemental Table 12. Due to non-convergence, indirect effects for the sex-stratified models were not bootstrapped. Most effects were similar between child sex, with the exception of the association between maternal depression and harsh parenting, as shown in previous models. Adjusting for assessors did not change the results (see Supplemental Table 13). In addition, sensitivity analyses of missing data on IPV and ACEs did not show different results (presented in Supplemental Table 14).

## Discussion

This study investigated whether maternal warmth, stimulation, and harsh parenting mediated associations between maternal depression and child mental health, socioemotional, and cognitive skills in a sample of mothers and children from rural Pakistan. Our study is one of the first to examine longitudinal mediation of maternal depression by parenting domains within a LMIC. Overall, this study found evidence that maternal depression during infancy has a direct association with child mental health difficulties and socioemotional development at 36 months. There was also evidence that maternal depression at 12 months is associated with increased harsh parenting behaviors at 24 months. However, for most outcomes, there was no evidence that parenting behaviors were associated with child outcomes, nor were there indirect effects from maternal depression on child outcomes through parenting behaviors. The only exception was fine motor skills; we found that increased harsh parenting predicted decreased fine motor skills, and that there was an indirect effect of maternal depression on fine motor skills through harsh parenting.

The associations between maternal depression and increased mental health difficulties and socioemotional concerns are consistent with previous research showing intergenerational transmission of mental health difficulties from mothers to offspring (Goodman et al., 2011). We did not find direct effects of maternal depression on child receptive language or fine motor skills. It is possible that other environmental factors, including poverty, violence exposure, nutrition, and exposure to disease are the main drivers of child cognitive outcomes, overshadowing any impact of maternal depression. It may be that these factors are common prior causes of both maternal depression and child cognitive development and the relative contribution of maternal depression to child outcomes, after controlling for many of these factors, is relatively small.

Indeed, research has shown that child development is influenced by a myriad of factors beyond maternal psychopathology (Haft & Hoeft, 2017; Jeong et al., 2020; Kieling et al., 2011). In LMICs, low access to financial and healthcare resources may further exacerbate the effect of contextual risk factors on child development. It is also possible that there are unique protective factors in this context that reduce associations between maternal depression and child cognitive outcomes. For example, household characteristics, such as the presence of extended family, may serve an extra support for both mothers and children, thereby buffering some of the harmful effects of maternal depression (Chung et al., 2020). Our sample was not large enough to compare these associations between nuclear family households and extended family households, although this may be an important consideration in future research.

We found that maternal depression at 12 months predicted increased harsh parenting behaviors at 24 months, aligning with previous research (Wolford et al., 2019). Depression may interfere with a mother’s ability to practice emotion regulation and show patience with her child, resulting in increased harsh parenting behaviors, such as negative statements or physical punishment (Dix & Meunier, 2009). This association was stronger in girls compared to boys.

This points to potential differences in how maternal depression influences parenting behaviors based on child sex. Previous research has suggested that parents show differences in their discipline strategies and play interactions based on child sex (McKee & Olson, 2007; Morawska, 2020). In Pakistan in particular, mothers report that they are more likely to show warmth to their girls compared to boys and are more likely to encourage autonomy when interacting with their boys compared to girls (Stewart et al., 2006). Our results suggest that maternal depression may influence these sex-specific maternal-child interactions, such that mothers with depression are more likely to show negative behaviors (i.e., negative affect, negative verbal statements) when interacting with their female children. Future research could further explore how maternal depression, parenting behaviors, and child sex interact to influence developmental outcomes.

We did not find consistent associations between parenting behaviors and child outcomes in this study, which is contrary to some previous research in other settings (Black et al., 2007; Jeong et al., 2016; Urke et al., 2018). This suggests that, in this sample, parenting at 24 months postpartum is not a strong mechanism linking maternal depression at 12 months to child outcomes at 36 months. It may be true that parenting mediates the relationship between maternal depression and child outcomes through different parenting domains than the ones this study explored. For instance, our measure of parenting was gathered in the context of a play task, which does not necessarily elicit mothers’ discipline strategies or their responses to child misbehavior. This may be an important facet of parenting behavior to examine in future studies, as mothers may show more variation in both warmth and harshness while responding to challenging child behavior (e.g., disobedience, tantrums). Indeed, past research has shown that mothers with depression are more likely to engage in punitive discipline strategies, such as corporal punishment, in response to child disobedience compared to mothers without depression (McLearn et al., 2006).

It is also possible that other, non-parenting related, pathways from maternal depression to child development have stronger influence in this context. For instance, maternal depression may influence children’s developing stress response systems, altering their growth and brain development, which then may place them at increased risk for poor socioemotional and cognitive outcomes (Van Den Bergh et al., 2017). There may also be other biological pathways linking maternal depression and child development, including epigenetic regulation, microbiome alterations, and exposure to infection early in life (Herba et al., 2016).

We found some evidence that harsh parenting at 24 months predicts lower fine motor skills at 36 months. Previous research has suggested that harsh parenting is detrimental to children’s cognitive development more generally (Berthelon et al., 2020). Mothers who struggle with harsh parenting may also be less likely to encourage their young children to practice their fine motor skills through manipulating small objects or attempting complex motor tasks. Our results suggest that there is an indirect effect of maternal depression on fine motor skills through harsh parenting. This suggests that, although there is not a direct effect of maternal depression on fine motor skill in this sample, harsh parenting may be a mechanism conferring risk for suboptimal cognitive development among children whose mothers struggle with depression (Goodman et al., 2020).

This study had many strengths, including the use of longitudinal data spanning from pregnancy to child age 36 months, a large population-representative sample, and developmental outcomes across several domains. In addition, this study uses an observational measure of parenting, allowing us to gain a nuanced understanding of parenting behavior and reduce the bias inherent in self-reported parenting measures. Finally, this study includes a sample of mothers and children in rural Pakistan, which provides much-needed evidence on the associations among maternal depression and child development in a low-resource, South Asian context.

This study also includes important limitations. The mental health and socioemotional development domains were mother-reported. Future studies would benefit from including multi-informant and observational measures of child behavior, especially as it relates to mental health and socioemotional skills. Additionally, the current analyses did not account for potential bidirectional effects between maternal depression and child behavior, as it was beyond the scope of this manuscript. Several studies have demonstrated that maternal depression and child behaviors, such as aggression, influence on another over time (Allen et al., 2019; Kuckertz et al., 2018).

This study showed small or null indirect effects in the associations between maternal depression and child development through parenting behaviors in a sample of mother-child dyads in rural Pakistan. The findings point to the importance of investigating alternative pathways between maternal depression and child development (e.g., the role of extended family, stress system functioning). Future studies may also focus on child, parent, and family characteristics when disentangling the complex associations among maternal mental health, parenting, and child health. Results suggest that parenting may be one of many targets for supporting child development and mitigating the effects of maternal depression among families in a low-resource, South Asian context.

## Data Availability

All data produced in the present study are available upon reasonable request to the authors.

## Acknowledgements

We would like to thank the team at the Human Development Research Foundation (HDRF) including Rakshanda Liaqat, Tayyiba Abbasi, Maria Sharif, Samina Bilal, Anum Nisar, Amina Bibi, Shaffaq Zufiqar, Ahmed Zaidi, Ikhlaq Ahmad, and Najia Atif. Finally, we are very grateful to the families who participated in the study.

## Notes

### Competing Interest Statement

The authors have declared no competing interest.

### Funding Statement

This study was funded by the National Institute of Mental Health (U19MH95687) and National Institute of Child Health and Development (R01 HD075875).

### Author Declarations

The Institutional Review Board of the University of North Carolina at Chapel Hill gave ethical approval for this work.

