## Appendix for "Longitudinal pathways between maternal depression, parenting behaviors, and early childhood development: a mediation analysis"

Appendix A: Items included in the parenting, child mental health, and child socioemotional development factors

**Parental Warmth**

OMCI 4: Mom shows positive touch

OMCI 6: Mom makes positive verbal statements

OMCI 9: Moms is sensitive to child’s needs, play is child-centered

**Parental Stimulation**

OMCI 10: Mom points to objects in book

OMCI 11: Mom questions child

**Harsh Parenting**

OMCI 1: Mom shows positive affect towards child

OMCI 2: Mom shows negative affect towards child

OMCI 5: Mom shows negative touch

OMCI 7: Mom makes negative verbal statements

OMCI 8: Mom shows intrusiveness

**Mental health difficulties: Strengths and Difficulties Questionnaire (SDQ)***

SDQ 2: Restless, overactive, cannot stay still for long

SDQ 3: Often complains of headaches, stomachaches, or sickness

SDQ 5: Often loses temper

SDQ 7: Generally well-behaved

SDQ 10: Constantly fidgeting or squirming

SDQ 11: Has at least one good friend

SDQ 12: Often fights with other children or bullies them

SDQ 13: Often unhappy, depressed, or tearful

SDQ 16: Nervous or clingy in new situations, easily loses confidence

SDQ 19: Picked on or bullied by other children

SDQ 23: Gets along better with adults than with other children

SDQ 24: Many fears, easily scared

SDQ 25: Good attention span, sees work through to the end

**Socioemotional development: Ages and Stages Questionnaire- Socioemotional (ASQ)****

ASQ 5: When upset, can your child calm down within 15 minutes?

ASQ 6: Does your child seem too friendly with strangers?

ASQ 7: Can your child settle herself down after a period of exciting activity?

ASQ 8: Can you child move from one activity to the next with little difficulty?

ASQ 9: Does your child seem happy?

ASQ 10: Is your child interested in things around him?

ASQ 11: Does your child do what you ask her to do?

ASQ 12: Does your child seem more active than other children her age?

ASQ 13: Can your child stay with the activities she enjoys for at least five minutes?

ASQ 14: Do you and your child enjoy mealtimes together?

ASQ 15: Does your child have eating problems?

ASQ 18: Does your child follow routine directions?

ASQ 20: Does your child check to make sure you are near when exploring new places, such as market or relative home?

ASQ 23: Does your child stay away from dangerous things, such as fire and moving cars?

ASQ 24: Does your child destroy or damage things on purpose?

ASQ 25: Does your child use words to describe her feelings and the feelings of others?

ASQ 26: Does your child name his/her age fellow in neighborhood?

ASQ 30: Has anyone expressed concerns about your child’s behaviours?

*SDQ items 6, 8, 9, 15, 18, 21, and 22 were removed due to redundancy. SDQ items 1, 4, 14, 17, 20 were part of the prosocial scale, so were not included in the mental health difficulties score.

**ASQ items 1, 2, 3, 4, 16, 17, 19, 21, 22, 27, 28, and 29 were removed due to redundancy with other items or because they showed low variability.

Supplemental Table 1.

Mediation model with maternal depression, parenting, and child mental health difficulties stratified by child sex

| Path | Female | Male |
| --- | --- | --- |
|  | β (95% CI) | β (95% CI) |
| SCID on SDQ | 0.37 (0.08, 0.65) | 0.39 (0.08, 0.70) |
| Harsh Parenting on SDQ | 0.05 (-0.11, 0.21) | -0.01 (-0.17, 0.14) |
| Warmth on SDQ | -0.09 (-0.38, 0.21) | -0.00 (-0.16, 0.15) |
| Stimulation on SDQ | -0.02 (-0.25, 0.21) | 0.06 (-0.16, 0.27) |
| SCID on Harsh Parenting | 0.50 (0.08, 0.92) | 0.30 (-0.75, 1.34) |
| SCID on Warmth | -0.20 (-0.87, 0.46) | 0.09 (-0.22, 0.42) |
| SCID on Stimulation | -0.03 (-0.41, 0.35) | -0.37 (-0.85, 0.11) |
| Indirect Effect of SCID on SDQ via Harsh Parenting | 0.025 (-0.03, 0.10) | -0.003 (-0.06, 0.08) |
| Indirect Effect of SCID on SDQ via Warmth | 0.027 (-0.03, 0.06) | 0.000 (-0.04, 0.03) |
| Indirect Effect of SCID on SDQ via Stimulation | 0.001 (-0.02, 0.04) | -0.021 (-0.14, 0.02) |

*Notes:* SCID = Structured Clinical Interview for the DSM-IV (binary maternal depression measure), SDQ = Strengths and Difficulties Questionnaire. Confidence intervals for indirect effects are bootstrapped.

Supplemental Table 2.

Mediation models with maternal depression predicting child mental health difficulties (subscales of the SDQ) through parenting behaviors

| Path | Emotional Problems | Conduct Problems | Hyperactivity | Peer Problems | Prosocial Behavior |
| --- | --- | --- | --- | --- | --- |
|  | β (95% CI) | β (95% CI) | β (95% CI) | β (95% CI) | β (95% CI) |
| SCID on SDQ | 0.34  (0.11, 0.56) | 0.39  (0.18, 0.60) | 0.13  (-0.19, 0.45) | 0.22  (0.04, 0.40) | 0.05  (-0.23, 0.33) |
| Harsh Parenting on SDQ | 0.01  (-0.10, 0.12) | 0.02  (-0.10, 0.14) | -0.05  (-0.21, 0.11) | 0.04  (-0.10, 0.18) | -0.09  (-0.29, 0.12) |
| Warmth on SDQ | -0.11  (-0.21, -0.01) | -0.08  (-0.17, 0.02) | 0.02  (-0.11, 0.16) | -0.02  (-0.10, 0.06) | -0.03  (-0.18, 0.11) |
| Stimulation on SDQ | 0.07  (-0.03, 0.16) | 0.07  (-0.03, 0.16) | 0.01  (-0.16, 0.18) | 0.02  (-0.07, 0.11) | -0.03  (-0.15, 0.08) |
| SCID on Harsh Parenting | 0.43  (0.15, 0.71) | 0.43  (0.15, 0.71) | 0.43  (0.15, 0.71) | 0.43  (0.15, 0.71) | 0.43  (0.15, 0.71) |
| SCID on Warmth | -0.07  (-0.28, 0.15) | -0.07  (-0.29, 0.15) | -0.07  (-0.29, 0.15) | -0.07  (-0.29, 0.15) | -0.07  (-0.29, 0.15) |
| SCID on Stimulation | -0.18  (-0.44, 0.08) | -0.18  (-0.45, 0.10) | -0.18  (-0.50, 0.15) | -0.18  (-0.44, 0.08) | -0.18  (-0.44, 0.08) |
| Indirect Effect of SCID on SDQ through HP | 0.004  (-0.03, 0.05) | 0.008  (-0.04, 0.06) | -0.022  (-0.07, 0.03) | 0.019  (-0.02, 0.07) | -0.038  (-0.10, 0.02) |
| Indirect Effect of SCID on SDQ through Warmth | 0.008  (-0.02, 0.04) | 0.005  (-0.01, 0.03) | -0.002  (-0.02, 0.01) | 0.001  (-0.01, 0.02) | 0.002  (-0.02, 0.03) |
| Indirect Effect of SCID on SDQ through Stimulation | -0.011  (-0.04, 0.01) | -0.012  (-0.05, 0.01) | -0.002  (-0.05, 0.03) | -0.003  (-0.03, 0.01) | 0.006  (-0.04, 0.02) |

Notes: SCID = Structured Clinical Interview for the DSM-IV (binary maternal depression measure), SDQ = Strengths and Difficulties Questionnaire. Due to convergence issues, the Peer Problems scale is modeled as a manifest variable (sum of items). No items were excluded from any scales. Higher scores on all scales indicate increased mental health difficulties, except for Prosocial Behavior (on which higher scores indicate increased prosocial behaviors). Confidence intervals for indirect effects are bootstrapped.

Supplemental Table 3.

Mediation model with maternal depression predicting child mental health through parenting behaviors, adjusted for assessors at 12, 24, and 36 months

| Path | β (95% CI) |
| --- | --- |
| SCID on SDQ | 0.34 (0.19, 0.49) |
| Harsh Parenting on SDQ | 0.05 (-0.04, 0.14) |
| Warmth on SDQ | -0.03 (-0.1, 0.05) |
| Stimulation on SDQ | 0 (-0.07, 0.07) |
| SCID on Harsh Parenting | 0.4 (0.07, 0.72) |
| SCID on Warmth | -0.23 (-0.44, -0.02) |
| SCID on Stimulation | -0.2 (-0.43, 0.02) |
| Indirect Effect of SCID on SDQ through HP | 0.02 (-0.01, 0.07) |
| Indirect Effect of SCID on SDQ through Warmth | 0.01 (-0.01, 0.03) |
| Indirect Effect of SCID on SDQ through Stimulation | 0 (-0.02, 0.02) |

Notes: SCID = Structured Clinical Interview for the DSM-IV (binary maternal depression measure), SDQ = Strengths and Difficulties Questionnaire. In this model, we adjust for the 12-month assessor when regressing on the SCID, the 24-month assessor when regressing on the parenting mediators, and the 36-month assessor when regressing on the outcome.

Supplemental Table 4.

Sensitivity Analysis on Missingness in IPV and ACES with the SDQ outcome.

| Path |  | “Best Case” Imputation | “Worst Case” Imputation |
| --- | --- | --- | --- |
| SCID on SDQ |  | 0.407 (0.223, 0.592) | 0.418 (0.233, 0.602) |
| Harsh Parenting on SDQ |  | -0.005 (-0.107, 0.098) | -0.006 (-0.103, 0.091) |
| Warmth on SDQ |  | -0.034 (-0.129, 0.06) | -0.035 (-0.129, 0.06) |
| Stimulation on SDQ |  | -0.002 (-0.1, 0.097) | -0.001 (-0.099, 0.097) |
| SCID on Harsh Parenting |  | 0.357 (0.085, 0.628) | 0.361 (0.092, 0.63) |
| SCID on Warmth |  | -0.112 (-0.317, 0.092) | -0.106 (-0.31, 0.097) |
| SCID on Stimulation |  | -0.225 (-0.53, 0.081) | -0.231 (-0.547, 0.085) |
| Indirect Effect of SCID on SDQ through HP |  | -0.002 (-0.032, 0.037) | -0.002 (-0.034, 0.037) |
| Indirect Effect of SCID on SDQ through Warmth |  | 0.004 (-0.012, 0.025) | 0.004 (-0.012, 0.023) |
| Indirect Effect of SCID on SDQ through Stimulation |  | 0 (-0.026, 0.027) | 0 (-0.027, 0.027) |

Notes: SCID = Structured Clinical Interview for the DSM-IV (binary maternal depression measure), SDQ = Strengths and Difficulties Questionnaire. In “Best Case” imputation, all women with missing data on IPV were assigned a zero (i.e., no IPV experienced) and all women with missing ACEs data were assigned zero (i.e., no adverse childhood experiences). In “Worst Case” imputation, all women with missing IPV data were assigned a one (i.e., experienced IPV) and all women with missing ACEs data were assigned the median ACEs score (Median = 1.00)

Supplemental Table 5.

Mediation model with maternal depression, parenting, and child socioemotional development stratified by child sex

| Path | Female | Male |
| --- | --- | --- |
|  | β (95% CI) | β (95% CI) |
| SCID on ASQ | 0.16 (-0.12, 0.44) | 0.40 (0.13, 0.68) |
| Harsh Parenting on ASQ | -0.02 (-0.15, 0.11) | 0.01 (-0.11, 0.13) |
| Warmth on ASQ | -0.10 (-0.24, 0.04) | -0.07 (-0.21, 0.06) |
| Stimulation on ASQ | -0.04 (-0.16, 0.08) | -0.02 (-0.15, 0.11) |
| SCID on Harsh Parenting | 0.50 (0.23, 0.77) | 0.30 (0.01, 0.59) |
| SCID on Warmth | -0.22 (-0.51, 0.07) | 0.09 (-0.21, 0.40) |
| SCID on Stimulation | -0.03 (-0.29, 0.23) | -0.38 (-0.66, -0.09) |
| Indirect Effect of SCID on ASQ via Harsh Parenting | -0.01 (-0.08, 0.06) | 0.004 (-0.03, 0.04) |
| Indirect Effect of SCID on ASQ via Warmth | 0.022 (-0.02, 0.06) | -0.007 (-0.03, 0.02) |
| Indirect Effect of SCID on ASQ via Stimulation | 0.001 (-0.01, 0.01) | 0.007 (-0.04, 0.06) |

*Notes*: SCID = Structured Clinical Interview for the DSM-IV (binary maternal depression measure), ASQ = Ages and Stages Questionnaire.

Supplemental Table 6.

Mediation model with maternal depression predicting child socioemotional development (modeled as a manifest variable) through parenting behaviors

| Path | β (95% CI) |
| --- | --- |
| SCID on ASQ | 0.26 (0.08, 0.44) |
| Harsh Parenting on ASQ | 0.09 (0.00, 0.17) |
| Warmth on ASQ | -0.07 (-0.16, 0.02) |
| Stimulation on ASQ | -0.05 (-0.13, 0.03) |
| SCID on Harsh Parenting | 0.44 (0.23, 0.64) |
| SCID on Warmth | -0.08 (-0.29, 0.13) |
| SCID on Stimulation | -0.18 (-0.38, 0.01) |
| Indirect Effect of SCID on ASQ through HP | 0.04 (-0.00, 0.08) |
| Indirect Effect of SCID on ASQ through Warmth | 0.01 (-0.01, 0.02) |
| Indirect Effect of SCID on ASQ through Stimulation | 0.01 (-0.01, 0.03) |

Notes: SCID = Structured Clinical Interview for the DSM-IV (binary maternal depression measure), ASQ = Ages and Stages Questionnaire, modeled as a manifest variable representing the sum score of ASQ items.

Supplemental Table 7.

Mediation model with maternal depression predicting child socioemotional development through parenting behaviors, adjusted for assessors at 12, 24, and 36 months

| Path | β (95% CI) |
| --- | --- |
| SCID on ASQ | 0.22 (0.04, 0.4) |
| Harsh Parenting on ASQ | 0.02 (-0.06, 0.1) |
| Warmth on ASQ | -0.07 (-0.16, 0.01) |
| Stimulation on ASQ | -0.02 (-0.1, 0.06) |
| SCID on Harsh Parenting | 0.4 (0.19, 0.61) |
| SCID on Warmth | -0.23 (-0.43, -0.03) |
| SCID on Stimulation | -0.2 (-0.39, -0.01) |
| Indirect Effect of SCID on ASQ through HP | 0.01 (-0.02, 0.04) |
| Indirect Effect of SCID on ASQ through Warmth | 0.02 (-0.01, 0.04) |
| Indirect Effect of SCID on ASQ through Stimulation | 0 (-0.01, 0.02) |

Notes: SCID = Structured Clinical Interview for the DSM-IV (binary maternal depression measure), ASQ = Ages and Stages Questionnaire. In this model, we adjust for the 12-month assessor when regressing on the SCID, the 24-month assessor when regressing on the parenting mediators, and the 36-month assessor when regressing on the outcome.

Supplemental Table 8.

Sensitivity Analysis on Missingness in IPV and ACES with the ASQ outcome

| Path | “Best Case” Imputation | “Worst Case” Imputation |
| --- | --- | --- |
| SCID on ASQ | 0.261 (0.072, 0.451) | 0.268 (0.078, 0.457) |
| Harsh Parenting on ASQ | 0.009 (-0.086, 0.105) | 0.009 (-0.086, 0.105) |
| Warmth on ASQ | -0.08 (-0.178, 0.019) | -0.079 (-0.178, 0.019) |
| Stimulation on ASQ | -0.022 (-0.113, 0.068) | -0.022 (-0.113, 0.068) |
| SCID on Harsh Parenting | 0.366 (0.166, 0.565) | 0.361 (0.161, 0.561) |
| SCID on Warmth | -0.113 (-0.316, 0.091) | -0.119 (-0.323, 0.085) |
| SCID on Stimulation | -0.229 (-0.421, -0.037) | -0.222 (-0.414, -0.029) |
| Indirect Effect of SCID on ASQ through HP | 0.003 (-0.032, 0.039) | 0.003 (-0.031, 0.038) |
| Indirect Effect of SCID on ASQ through Warmth | 0.009 (-0.011, 0.029) | 0.009 (-0.011, 0.03) |
| Indirect Effect of SCID on ASQ through Stimulation | 0.005 (-0.016, 0.026) | 0.005 (-0.016, 0.026) |

Notes: SCID = Structured Clinical Interview for the DSM-IV (binary maternal depression measure), ASQ = Ages and Stages Questionnaire. In “Best Case” imputation, all women with missing data on IPV were assigned a zero (i.e., no IPV experienced) and all women with missing ACEs data were assigned zero (i.e., no adverse childhood experiences). In “Worst Case” imputation, all women with missing IPV data were assigned a one (i.e., experienced IPV) and all women with missing ACEs data were assigned the median ACEs score (Median = 1.00)

Supplemental Table 9.

Mediation model with maternal depression, parenting, and child receptive language stratified by child sex

| Path | Female | Male |
| --- | --- | --- |
|  | β (95% CI) | β (95% CI) |
| SCID on Receptive Language | -0.01 (-0.25, 0.24) | -0.25 (-0.51, 0.01) |
| Harsh Parenting on Receptive Language | -0.05 (-0.16, 0.07) | -0.01 (-0.11, 0.10) |
| Warmth on Receptive Language | 0.12 (0.00, 0.23) | 0 (-0.12, 0.12) |
| Stimulation on Receptive Language | 0.01 (-0.10, 0.12) | 0.13 (0.00, 0.26) |
| SCID on Harsh Parenting | 0.50 (0.23, 0.77) | 0.30 (0.01, 0.59) |
| SCID on Warmth | -0.21 (-0.50, 0.07) | 0.10 (-0.21, 0.41) |
| SCID on Stimulation | -0.03 (-0.30, 0.23) | -0.38 (-0.66, -0.09) |
| Indirect Effect of SCID on Receptive Language via Harsh Parenting | -0.02 (-0.08, 0.03) | 0.00 (-0.03, 0.03) |
| Indirect Effect of SCID on Receptive Language via Warmth | -0.02 (-0.07, 0.02) | 0.00 (-0.01, 0.01) |
| Indirect Effect of SCID on Receptive Language via Stimulation | 0.00 (0.00, 0.00) | -0.05 (-0.11, 0.01) |

*Notes*: SCID = Structured Clinical Interview for the DSM-IV (binary maternal depression measure)

Supplemental Table 10.

Mediation model with maternal depression predicting child receptive language through parenting behaviors, adjusted for assessors at 12, 24, and 36 months

| Path | β (95% CI) |
| --- | --- |
| SCID on Bayley’s Scaled Receptive Score | -0.12 (-0.28, 0.04) |
| Harsh Parenting on Bayley’s Scaled Receptive Score | -0.05 (-0.13, 0.02) |
| Warmth on Bayley’s Scaled Receptive Score | 0.08 (0.01, 0.16) |
| Stimulation on Bayley’s Scaled Receptive Score | 0.04 (-0.04, 0.11) |
| SCID on Harsh Parenting | 0.4 (0.19, 0.61) |
| SCID on Warmth | -0.23 (-0.43, -0.03) |
| SCID on Stimulation | -0.2 (-0.39, -0.01) |
| Indirect Effect of SCID on Bayley’s Scaled Receptive Score through HP | -0.02 (-0.05, 0.01) |
| Indirect Effect of SCID on Bayley’s Scaled Receptive Score through Warmth | -0.02 (-0.04, 0.01) |
| Indirect Effect of SCID on Bayley’s Scaled Receptive Score through Stimulation | -0.01 (-0.02, 0.01) |

Notes: SCID = Structured Clinical Interview for the DSM-IV (binary maternal depression measure). In this model, we adjust for the 12-month assessor when regressing on the SCID, the 24-month assessor when regressing on the parenting mediators, and the 36-month assessor when regressing on the outcome.

Supplemental Table 11.

Sensitivity analysis on missingness in IPV and ACEs with Receptive Language outcome

| Path | “Best Case” Imputation | “Worst Case” Imputation |
| --- | --- | --- |
| SCID on Bayley’s Scaled Receptive Score | -0.11 (-0.289, 0.069) | -0.132 (-0.311, 0.047) |
| Harsh Parenting on Bayley’s Scaled Receptive Score | -0.016 (-0.097, 0.066) | -0.014 (-0.096, 0.068) |
| Warmth on Bayley’s Scaled Receptive Score | 0.061 (-0.023, 0.145) | 0.061 (-0.023, 0.145) |
| Stimulation on Bayley’s Scaled Receptive Score | 0.046 (-0.037, 0.129) | 0.046 (-0.037, 0.129) |
| SCID on Harsh Parenting | 0.365 (0.165, 0.564) | 0.359 (0.159, 0.56) |
| SCID on Warmth | -0.11 (-0.313, 0.093) | -0.116 (-0.32, 0.088) |
| SCID on Stimulation | -0.229 (-0.42, -0.037) | -0.221 (-0.414, -0.029) |
| Indirect Effect of SCID on Bayley’s Scaled Receptive Score through HP | -0.006 (-0.036, 0.024) | -0.005 (-0.035, 0.024) |
| Indirect Effect of SCID on Bayley’s Scaled Receptive Score through Warmth | -0.007 (-0.022, 0.009) | -0.007 (-0.023, 0.009) |
| Indirect Effect of SCID on Bayley’s Scaled Receptive Score through Stimulation | -0.011 (-0.032, 0.01) | -0.01 (-0.031, 0.01) |

Notes: SCID = Structured Clinical Interview for the DSM-IV (binary maternal depression measure), In “Best Case” imputation, all women with missing data on IPV were assigned a zero (i.e., no IPV experienced) and all women with missing ACEs data were assigned zero (i.e., no adverse childhood experiences). In “Worst Case” imputation, all women with missing IPV data were assigned a one (i.e., experienced IPV) and all women with missing ACEs data were assigned the median ACEs score (Median = 1.00)

Supplemental Table 12.

Mediation model with maternal depression, parenting, and child fine motor skills stratified by child sex

| Path | Female | Male |
| --- | --- | --- |
|  | β (95% CI) | β (95% CI) |
| SCID on Fine Motor Skills | 0.13 (-0.11, 0.38) | 0.02 (-0.25, 0.28) |
| Harsh Parenting on Fine Motor Skills | -0.12 (-0.24, -0.01) | -0.07 (-0.19, 0.04) |
| Warmth on Fine Motor Skills | 0.07 (-0.04, 0.19) | -0.01 (-0.14, 0.12) |
| Stimulation on Fine Motor Skills | -0.04 (-0.14, 0.07) | 0.03 (-0.1, 0.16) |
| SCID on Harsh Parenting | 0.51 (0.24, 0.78) | 0.31 (0.02, 0.61) |
| SCID on Warmth | -0.22 (-0.5, 0.07) | 0.10 (-0.21, 0.41) |
| SCID on Stimulation | -0.03 (-0.3, 0.23) | -0.37 (-0.66, -0.09) |
| Indirect Effect of SCID on Fine Motor Skills via Harsh Parenting | -0.06 (-0.13, 0.00) | -0.02 (-0.07, 0.02) |
| Indirect Effect of SCID on Fine Motor Skills via Warmth | -0.02 (-0.05, 0.02) | 0.00 (-0.01, 0.01) |
| Indirect Effect of SCID on Fine Motor Skills via Stimulation | 0.00 (-0.01, 0.01) | -0.01 (-0.06, 0.04) |

*Notes*: SCID = Structured Clinical Interview for the DSM-IV (binary maternal depression measure)

Supplemental Table 13.

Mediation model with maternal depression predicting child fine motor skills through parenting behaviors, adjusted for assessors at 12, 24, and 36 months

| Path | β (95% CI) |
| --- | --- |
| SCID on Bayley’s Scaled Fine Motor Score | 0.08 (-0.09, 0.25) |
| Harsh Parenting on Bayley’s Scaled Fine Motor Score | -0.12 (-0.22, -0.01) |
| Warmth on Bayley’s Scaled Fine Motor Score | 0.04 (-0.05, 0.13) |
| Stimulation on Bayley’s Scaled Fine Motor Score | -0.02 (-0.09, 0.06) |
| SCID on Harsh Parenting | 0.4 (0.09, 0.72) |
| SCID on Warmth | -0.23 (-0.44, -0.03) |
| SCID on Stimulation | -0.2 (-0.43, 0.02) |
| Indirect Effect of SCID on Bayley’s Scaled Fine Motor Score through HP | -0.05 (-0.1, -0.01) |
| Indirect Effect of SCID on Bayley’s Scaled Fine Motor Score through Warmth | -0.01 (-0.04, 0.01) |
| Indirect Effect of SCID on Bayley’s Scaled Fine Motor Score through Stimulation | 0 (-0.02, 0.02) |

Notes: Notes: SCID = Structured Clinical Interview for the DSM-IV (binary maternal depression measure). In this model, we adjust for the 12-month assessor when regressing on the SCID, the 24-month assessor when regressing on the parenting mediators, and the 36-month assessor when regressing on the outcome.

Supplemental Table 14.

Sensitivity analysis on missingness in IPV and ACES with Fine Motor Skills outcome

| Path | “Best Case” Imputation | “Worst Case” Imputation |
| --- | --- | --- |
| SCID on Bayley’s Scaled Fine Motor Score | 0.089 (-0.099, 0.277) | 0.103 (-0.085, 0.29) |
| Harsh Parenting on Bayley’s Scaled Fine Motor Score | -0.106 (-0.207, -0.006) | -0.107 (-0.208, -0.007) |
| Warmth on Bayley’s Scaled Fine Motor Score | 0.036 (-0.05, 0.123) | 0.036 (-0.05, 0.122) |
| Stimulation on Bayley’s Scaled Fine Motor Score | -0.025 (-0.112, 0.063) | -0.025 (-0.112, 0.063) |
| SCID on Harsh Parenting | 0.371 (0.1, 0.641) | 0.376 (0.107, 0.645) |
| SCID on Warmth | -0.119 (-0.325, 0.087) | -0.112 (-0.318, 0.093) |
| SCID on Stimulation | -0.221 (-0.491, 0.048) | -0.228 (-0.498, 0.041) |
| Indirect Effect of SCID on Fine Motor through HP | -0.039 (-0.09, -0.005) | -0.04 (-0.09, -0.006) |
| Indirect Effect of SCID on Fine Motor through Warmth | -0.004 (-0.027, 0.009) | -0.004 (-0.026, 0.008) |
| Indirect Effect of SCID on Fine Motor through Stimulation | 0.006 (-0.019, 0.03) | 0.006 (-0.02, 0.03) |

Notes: SCID = Structured Clinical Interview for the DSM-IV (binary maternal depression measure), In “Best Case” imputation, all women with missing data on IPV were assigned a zero (i.e., no IPV experienced) and all women with missing ACEs data were assigned zero (i.e., no adverse childhood experiences). In “Worst Case” imputation, all women with missing IPV data were assigned a one (i.e., experienced IPV) and all women with missing ACEs data were assigned the median ACEs score (Median = 1.00)
